# Simultaneous Heart and Kidney Transplantation utilizing Circulatory Death Donors in the United States

**DOI:** 10.1101/2024.08.16.24312145

**Authors:** Sooyun C. Tavolacci, Kenji Okumura, Ameesh Isath, Gabriel B. Rodriguez, Corazon De La Pena, Junichi Shimamura, Steven L. Lansman, Suguru Ohira

## Abstract

**Objective:** Heart transplants utilizing donors from circulatory death (DCD) allografts are rapidly growing with the potential to expand the donor pool. However, little is known about the use of DCD donors for simultaneous heart and kidney transplants (SHKT) compared to SHKT using brain death donors (DBD).

**Methods:** From May 22, 2020, to September 30, 2023, 1,129 adult patients received SHKT (DCD, N=91 vs. DBD, N=1,038), identified using the United Network for Organ Sharing database, excluding other multi-organ transplants and re-transplants. A 1:3 ratio propensity score matching was performed using 17 recipient characteristics and 7 donor characteristics. A total of 91 DCD and 273 DBD matched cases were compared.

**Results:** In the unmatched cohort, DCD recipients were older (DCD: 60 vs. DBD: 58 years, p=0.03) and had a lower rate of dialysis at transplant (27% vs. 40%, p=0.03) and status 1 to 2 patients (43% vs. 72%, p<.001). Donors were younger (30 vs. 32 years, p=0.02) in the DCD group. In the matched cohort, kidney delayed graft function (27% vs. 22%, p=0.29) was comparable, as were recipient survival (p=0.19), heart graft survival (p=0.19), and kidney graft survival (p=0.17). In multivariate Cox proportional hazards analysis, donor type (DCD) was not associated with an increased risk of mortality (HR=1.69, 95% Cl 0.90-3.16, p=0.10). Sub-group analysis showed that survival and freedom from graft failures were comparable between different modes of DCD recovery. The centers performing both DCD- and DBD-SHKT showed significantly shorter waitlist days with comparable transplant outcomes compared to centers that only performed DBD-SHKT.

Conclusions

SHKT using DCD donors yields comparable survival and graft outcomes to those using DBD donors. These findings will guide treatment strategies for heart transplant candidates with kidney dysfunction, including the selection of donors and patients and safety net policy options.

## Introduction

The number of simultaneous heart and kidney transplants (SHKT) has continued to grow in the last 20 years due to a surge of end-stage heart failure patients with impaired renal function^1,2^. SHKT improves survival and quality of life in this population by implanting two functional organs from a single donor^1,3^. However, the utilization of two organs for one recipient may be controversial given the overall shortage of donors^4^, thereby limiting access to this life-saving therapy. Furthermore, the complexity of two major surgeries and peri-operative management may potentially increase early morbidity and mortality compared to heart transplant alone (OHT)^5^. Heart transplant candidates on chronic hemodialysis are often listed for SHKT, but OHT alone can lead to the recovery of renal function through improvement of cardio-renal syndrome in patients with borderline renal function^6^. The new kidney safety net policy allows OHT recipients who meet eligibility criteria for kidney transplant to receive additional priority on kidney matches within a year post-OHT.

The utilization of donor after circulatory death (DCD) in the United States through the utilization of thoracoabdominal normothermic regional perfusion (NRP) or ex-vivo perfusion known as direct procurement and perfusion (DPP), has allowed for the expansion of donor pool with promising outcomes^7,8^. However, the warm ischemic period prior to organ recovery in DCD donors in comparison to DBD recovery, can potentially impact organ function, especially in the setting of multi-organ transplant. To date, clinical evidence of SHKT utilizing DCD donors remains limited, including kidney graft outcomes. This study aims to analyze the outcomes of SKHT utilizing DCD vs DBD recovery, providing a comprehensive evaluation of the efficacy and safety of SHKT in DCD donor populations.

## Patients and Methods

### Study Population

The United Network for Organ Sharing (UNOS) database as of March 31, 2024, was queried from May 22, 2020, to September 30, 2023. A total of 1,129 SHKTs in adult patients, excluding other multi-organ transplants and re-transplants, were identified. Out of the 1,129, 1,038 patients received DBD-SHKT, and 91 patients received DCD-SHKT. A 1:3 ratio propensity score matching was performed: DCD (N=91) and DBD (N=273).

Two sub-group analyses were performed in the non-matched cohort: (i) the different modes of recovery method in DCD-SHKT: ex-vivo perfusion during transport after the heart is explanted, known as DPP, vs. NRP of donor with subsequent cold storage. The reported duration from brain death time to aortic clamp time was used to categorize the recovery method similar to previous studies^9–11^, which showed a bimodal distribution with the minor mode at 20 minutes **(Supplementary Figure S1)**. An interval of brain death time to aortic clamp time less than 20 minutes was considered DPP, and greater than 20 minutes was considered NRP.

This study was approved by the New York Medical College Institutional Review Board (IRB#14680).

### Endpoints and Definitions

All definitions of variables are described on the UNOS website (https://unos.org/data/data-collection/). Ischemic time is defined as time from cross clamp to reperfusion. The primary outcomes were one-year recipient and kidney graft survival. UNOS defines heart or kidney graft failure as follows: a recipient’s transplanted organ is removed (e.g., re-transplant), a recipient dies, or a recipient is placed on a chronic allograft support system (e.g., hemodialysis for kidney transplant). Secondary outcomes included the rate of stroke, acute rejection, dialysis, and kidney delayed graft function (KDGF), which was defined as the requirement of hemodialysis in the first week after kidney transplant. The estimated post-transplant survival (EPTS) score of kidney recipients was calculated using four factors: age, current diagnosis of diabetes, prior solid organ transplants, and time on dialysis^12^. The kidney donor profile index (KDPI) of donors was calculated using ten donor variables that influence donor organ quality: age, height, weight, cause of death, last serum creatinine, history of diabetes, hypertension, hepatitis C serostatus, ethnicity, and the distinction between donation after DCD versus DBD^13^. For both EPTS and KDPI, a lower value predicts a higher chance of kidney graft survival.

### Statistical Analysis

Baseline characteristics and outcomes were reported as median with interquartile range for continuous variables or number and percentage for categorical variables. Wilcoxon rank sum test, Pearson’s Chi-squared test, and Fisher’s exact test were used for group comparison depending on variable types and distributions. 1:3 propensity score matching with the nearest neighbor was performed. After matching, all covariates had a standardized difference of 10% or less, which was considered a threshold for ideal balance between the two cohorts **(Supplementary Figure S2)**.

Overall survival curves were derived using the Kaplan-Meier method; differences between cohorts were assessed using the log-rank test. Adjusted post-transplant survival was modeled using multivariate Cox proportional hazards regression. Factors identified as clinically relevant based on previous literature or clinical knowledge were included in the multivariable regression analysis. The results were expressed as hazard ratios (HR) with a 95% confidence interval (CI). All test were 2-tailed, and a p-value of less than 0.05 was considered significant. All statistical analyses were performed using RStudio 4.3.2.

## Results

### Baseline Characteristics

In the unmatched cohort **(Table 1),** the DCD group had older recipients (DCD, 60 [53-65.5] vs. DBD, 58 [50.3-64] years, p=0.03), lower prevalence of dialysis at the time of SHKT (27% vs. 40%, p=0.03), and a lower rate of status 1 to 2 patients (43% vs. 72%, p<0.001). Recipients EPTS score (DCD, 41 [28-51] % vs. DBD, 36 [22-50] %, p=0.04) was higher in the DCD group **(Supplemental Figure S3)**. Donors were younger (30 [24-35] vs. 32 [25-39] years, p=0.02), had more White individuals (74% vs. 59%, p=0.01), and had lower serum creatinine level (0.7 [0.6-1.0] vs. 0.9 [0.7-1.2] mg/dl, p<0.001) in the DCD group. The KDPI score was higher in the DCD group (22 [13-32] % vs. 18 [7-32] %, p=0.04) **(Supplemental Figure S3)**. The distance from donor to recipient hospital was longer in the DCD group (344 [153.5-581.5] vs. 231.5 [92-390.8] miles, p<0.001), as was heart ischemic time (4.9 [3.4-6.4] vs. 3.5 [2.9-4] hours, p<0.001), and kidney ischemic time (19.2 [13.7-24.8] vs. 16.3 [9.3-22] hours, p=0.002). Days on the waiting list were comparable for both heart (42 [9.5-160.5] vs. 33 [11-107.8] days, p=0.42) and kidney (37 [10-164] vs. 29 [10-91] days, p=0.15) transplants.

**Table 1.** Baseline Recipient, Donor, and Organ Procurement Characteristics of Unmatched and Matched Cohorts.

| Variable | DCD-SHKT, N = 91 | DBD-SHKT, N = 1,038 | p-value | Matched DBD-SHKT, | p-value |
| --- | --- | --- | --- | --- | --- |
| **Age** | 60.0 (53.0-65.5) | 58.0 (50.3-64.0) | 0.032 | 59.0 (52.0-64.0) | 0.36 |
| **Male** | 74 (81%) | 832 (80%) | 0.90 | 214 (78%) | 0.55 |
| **Race** |  |  | 0.56 |  | 0.81 |
| Non-White | 55 (60%) | 588 (57%) |  | 161 (59%) |  |
| White | 36 (40%) | 450 (43%) |  | 112 (41%) |  |
| **Diabetes** | 50 (55%) | 485 (47%) | 0.16 | 144 (53%) | 0.72 |
| **BMI** | 27.0 (24.0-31.6) | 27.2 (23.9-31.0) | 0.96 | 27.0 (23.8-30.4) | 0.60 |
| **Dialysis at transplant** | 25 (27%) | 415 (40%) | 0.025 | 98 (36%) | 0.14 |
| **History of smoking** | 36 (40%) | 413 (40%) | >0.99 | 107 (39%) | 0.95 |
| **Symptomatic cerebrovascular** | 8 (8.8%) | 96 (9.2%) | >0.99 | 26 (9.5%) | 0.84 |
| **Infection requiring IV drug** | 6 (6.6%) | 159 (15%) | 0.035 | 19 (7.0%) | 0.90 |
| **Status** |  |  | <0.001 |  | 0.47 |
| Status 1-2 | 39 (43%) | 751 (72%) |  | 129 (47%) |  |
| Status 3-6 | 52 (57%) | 287 (28%) |  | 144 (53%) |  |
| **Primary Diagnosis** |  |  | 0.54 |  | 0.50 |
| Ischemic CM | 28 (31%) | 359 (35%) |  | 74 (27%) |  |
| Non-Ischemic CM | 63 (69%) | 679 (65%) |  | 199 (73%) |  |
| **Durable LVAD** | 17 (19%) | 138 (13%) | 0.20 | 58 (21%) | 0.60 |
| **IABP** | 8 (8.8%) | 285 (27%) | <0.001 | 27 (9.9%) | 0.76 |
| **Impella** | 1 (1.1%) | 70 (6.7%) | 0.057 | 3 (1.1%) | >0.99 |
| **Ventilator** | 2 (2.2%) | 21 (2.0%) | >0.99 | 6 (2.2%) | >0.99 |
| **ECMO** | 5 (5.5%) | 58 (5.6%) | >0.99 | 16 (5.9%) | 0.90 |
| **History of cardiac surgery** | 31 (34%) | 346 (33%) | 0.98 | 92 (34%) | 0.95 |
| **Initial EPTS score** | 41 (28-51%) | 36 (22-50%) | 0.042 | 39 (26-54%) | 0.62 |
| **Recipient ABO** |  |  | 0.35 |  | 0.99 |
| A | 30 (33%) | 395 (38%) |  | 93 (34%) |  |
| AB | 4 (4.4%) | 56 (5.4%) |  | 12 (4.4%) |  |
| B | 12 (13%) | 174 (17%) |  | 33 (12%) |  |
| O | 45 (49%) | 413 (40%) |  | 135 (49%) |  |
| **Donor age** | 30.0 (24.0-35.0) | 32.0 (25.0-39.0) | 0.015 | 28.0 (21.0-37.0) | 0.64 |
| **Donor male** | 76 (84%) | 801 (77%) | 0.21 | 220 (81%) | 0.53 |
| **Donor race** |  |  | 0.010 |  | 0.95 |
| Non-White | 24 (26%) | 423 (41%) |  | 73 (27%) |  |
| White | 67 (74%) | 615 (59%) |  | 200 (73%) |  |
| **Donor BMI** | 27.1 (24.3-31.6) | 27.1 (23.6-31.1) | 0.52 | 26.2 (23.0-31.2) | 0.25 |
| **Donor Creatinine** | 0.7 (0.6-1.0) | 0.9 (0.7-1.2) | <0.001 | 0.8 (0.6-1.0) | 0.24 |
| **Donor Cause of Death** |  |  | 0.17 |  | 0.72 |
| Head Trauma | 46 (51%) | 441 (42%) |  | 132 (48%) |  |
| Other | 45 (49%) | 597 (58%) |  | 141 (52%) |  |
| **Donor LVEF** | 64.0 (60.0-67.0) | 60.0 (57.0-65.0) | 0.13 | 63.0 (60.0-66.0) | 0.85 |
| **Donor KDPI score** | 22 (13-32%) | 18 (7-32%) | 0.04 | 11 (4-24%) | <0.001 |
| **ABO match** |  |  | 0.82 |  | 0.11 |
| Identical | 79 (87%) | 886 (85%) |  | 252 (92%) |  |
| Compatible | 12 (13%) | 152 (15%) |  | 21 (7.7%) |  |
| **Heart Procurement Distance** | 344.0 (153.5-581.5) | 231.5 (92.0-390.8) | <0.001 | 212.0 (92.0-366.0) | <0.001 |
| **Heart Waiting list Time (days)** | 42.0 (9.5-160.5) | 33.0 (11.0-107.8) | 0.42 | 52.0 (14.0-188.0) | 0.26 |
| **Heart Ischemic Time (hours)** | 4.9 (3.4-6.4) | 3.5 (2.9-4.0) | <0.001 | 3.5 (2.9-4.0) | <0.001 |
| **Heart Machine Perfusion** | 53 / 91 (58%) | 44 / 1,038 (4.2%) | <0.001 | 8 / 273 (2.9%) | <0.001 |
| **Kidney Cold Ischemic Time** | 19.2 (13.7-24.8) | 16.3 (9.3-22.0) | 0.002 | 16.6 (9.9-21.5) |  |
| **Kidney Waiting list Time (days)** | 37.0 (10.0-164.0) | 29.0 (10.0-91.0) | 0.15 | 43.0 (13.0-131.3) |  |
| **Kidney Status at transplant** |  |  | >0.99 |  |  |
| Active | 79 / 81 (98%) | 928 / 952 (97%) |  | 239 / 244 (98%) |  |
| Temporarily Inactive | 2 / 81 (2.5%) | 24 / 952 (2.5%) |  | 5 / 244 (2.0%) |  |
| **Kidney Side, Left** | 62 / 81 (77%) | 642 / 954 (67%) | 0.11 | 170 / 245 (69%) |  |

\* BMI: Body Mass Index, CM: Cardiomyopathy, LVAD: Left Ventricular Assist Device, IABP: Intra-Aortic Balloon Pump, ECMO: Extracorporeal Membrane Oxygenation, and LVEF: Left Ventricular Ejection Fraction, KDPI: Kidney Donor Profile Index, and EPTS: Estimated Post-Transplant Survival
\* Heart ischemic time is defined as the period from donor death to reperfusion in the recipient, including both true ischemia time and ex vivo perfusion or normothermic regional perfusion time.

After propensity score matching, there was no difference in recipient and donor characteristics between DCD vs. DBD SHKT including dialysis and the listing status at the time of SHKT **(Table 1)**. The distance of procurement (344 [153.5-581.5] vs. 212 [92-366] miles, p<0.001), heart ischemic time (4.9 [3.4-6.4] vs. 3.5 [2.9-4] hours, p<0.001), and kidney cold ischemic time (19.2 [13.7-24.8] vs. 16.6 [9.9-21.5] hours, p=0.002) were longer in the DCD group.

### Transplant Outcomes

The incidence of KDGF was comparable in both non-matched (DCD, 27% vs. DBD, 26%, p = 0.68) and matched cohort (DCD, 27% vs. DBD, 22%, p=0.29) **(Table 2)** while creatinine level at discharge was higher in the DCD group (DCD, 1.6 [1.2-2.4] vs. DBD, 1.2 [0.9-1.7) mg/dL, p<0.001). There were no differences in the incidence of acute rejection, dialysis, stroke, pacemaker placement, or length of stay. Recipient survival (median follow-up: 366.5 [182.75-730] days, p=0.19), freedom from heart graft failure (p=0.19), and kidney graft failure (p=0.17) was similar in both non-matched and matched cohort **(Figure 1)**. In the matched group, one-year survival was 87% in the DCD-SHKT group and 90.8% in the DBD-SHKT group (p=0.37). In multivariate Cox proportional hazards analysis, donor type (DCD) was not associated with an increased risk of mortality (HR=1.69, 95% CI 0.90-3.16, p=0.10) **(Table 3)**. Recipient age, female recipient, and dialysis at the time of SHKT were identified as predictors for mortality.

**Figure 1.**
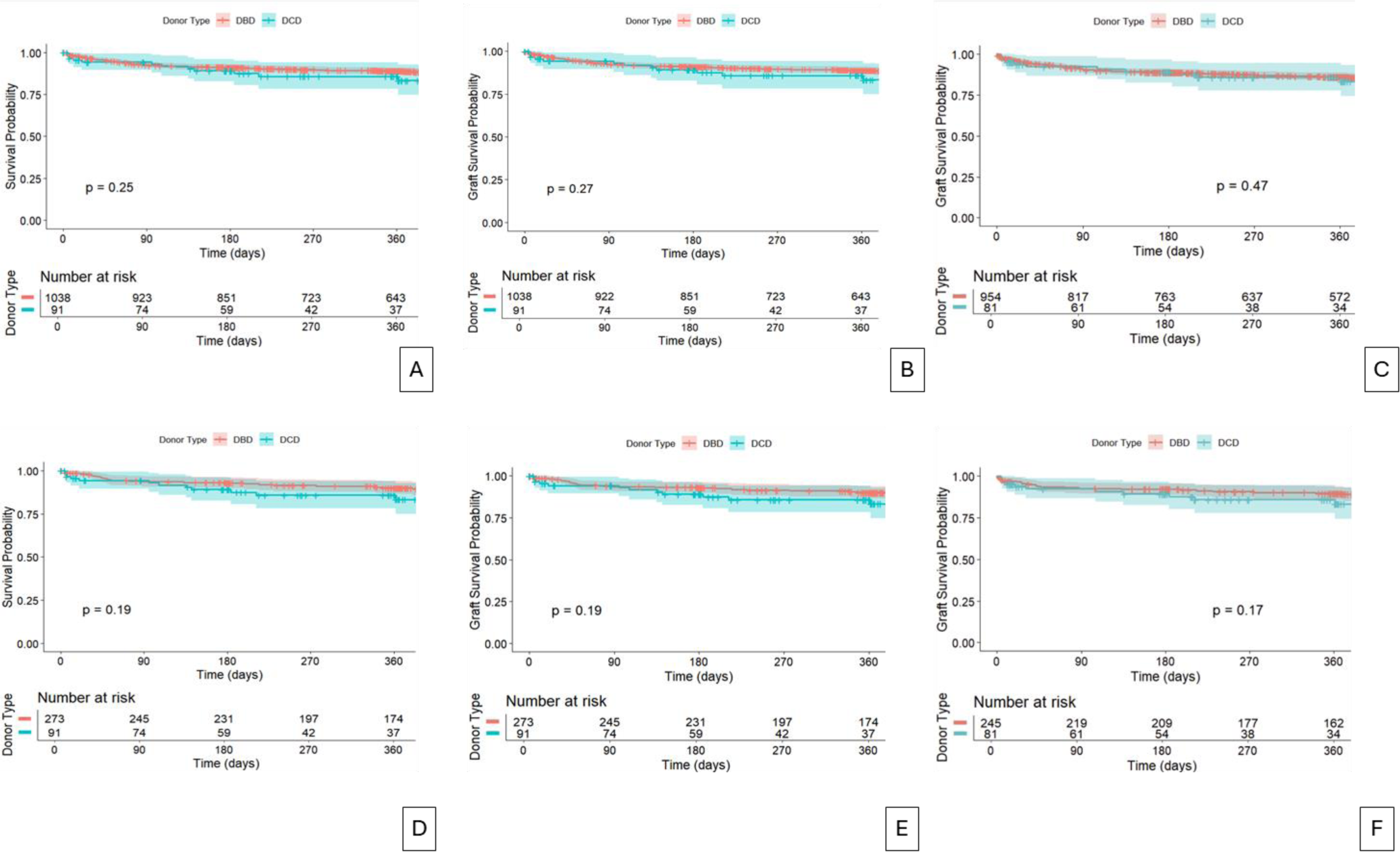
Recipient and Graft survivals outcome in Unmatched and Matched Cohorts. Unmatched cohort (A-C) and Matched cohort (D-F): A and D; Recipient survival, B and E; Heart graft survival, and C and F; Kidney graft survival

**Table 2.** Heart and Kidney Transplant Outcome of Unmatched and Matched Cohorts.

| Variable | DCD-SHKT, | DBD-SHKT, | p-value | Matched DBD-SHKT, | p-value |
| --- | --- | --- | --- | --- | --- |
|  | N = 91 | N = 1,038 |  | N = 273<sup>†</sup> |  |
| Acute Rejection | 6 (6.6%) | 89 (8.6%) | 0.65 | 14 (5.1%) | 0.79 |
| Dialysis | 34 (37%) | 365 (35%) | 0.77 | 85 (31%) | 0.33 |
| Stroke | 2 (2.2%) | 44 (4.3%) | 0.50 | 7 (2.6%) | >0.99 |
| Pacemaker | 2 (2.2%) | 14 (1.4%) | 0.85 | 3 (1.1%) | 0.79 |
| Heart Transplant Length of Stay | 20.0 (15.0-34.0) | 22.0 (15.0-33.0) | 0.73 | 21.0 (15.0-33.0) | 0.97 |
| Patient Mortality at 30 Days | 5 (5.5%) | 35 (3.4%) | 0.45 | 6 (2.2%) | 0.22 |
| Patient Mortality at 6 months | 9 (9.9%) | 91 (8.8%) | 0.87 | 19 (7.0%) | 0.50 |
| Patient Mortality at 1 year | 12 (13%) | 108 (10%) | 0.52 | 25 (9.2%) | 0.37 |
| Heart Graft Failure at 6 months | 9 (9.9%) | 92 (8.9%) | 0.89 | 19 (7.0%) | 0.50 |
| Heart Graft Failure at 1 year | 12 (13%) | 109 (11%) | 0.54 | 25 (9.2%) | 0.37 |
| Kidney Graft Failure at 6 months | 8 (9.9%) | 103 (11%) | 0.94 | 19 (7.8%) | 0.71 |
| Kidney Graft Failure at 1 year | 11 (14%) | 123 (13%) | >0.99 | 24 (9.8%) | 0.46 |
| Kidney Acute Rejection prior to discharge | 1 (1.2%) | 10 (1.1%) | >0.99 | 4 (1.7%) | >0.99 |
| Kidney Delayed Graft Function | 25 (27%) | 274 (26%) | 0.68 | 61 (22%) | 0.29 |
| Unknown | 10 (11%) | 84 (8.1%) |  | 28 (10%) |  |
| Resume Dialysis Maintenance after kidney transplant | 1 (1.1%) | 33 (3.2%) | 0.43 | 7 (2.6%) | 0.68 |
| Recipient creatinine at Discharge | 1.6 (1.2-2.4) | 1.3 (0.9-1.8) | <0.001 | 1.2 (0.9-1.7) | <0.001 |

**Table 3.** Multivariable Cox Proportional Hazards Analysis for Recipient Mortality.

|  | HR | Lower 95% CI | Upper 95% CI | p-value |
| --- | --- | --- | --- | --- |
| Donor Type (DCD) | 1.69 | 0.90 | 3.16 | 0.103 |
| Recipient Age | 1.02* | 1.01 | 1.04 | 0.012 |
| Recipient Gender (Female) | 1.76* | 1.20 | 2.57 | 0.004 |
| Recipient Ethnicity/Race (White) | 1.25 | 0.89 | 1.75 | 0.198 |
| Dialysis at transplant | 1.48* | 1.05 | 2.08 | 0.024 |
| IV antibiotics at transplant | 1.36 | 0.88 | 2.09 | 0.166 |
| Primary Diagnosis (Ischemic cardiomyopathy) | 1.21 | 0.85 | 1.70 | 0.288 |
| Durable LVAD (Yes) | 1.41 | 0.91 | 2.20 | 0.122 |
| IABP (No) | 1.17 | 0.77 | 1.78 | 0.474 |
| End Status (Status 1-2) | 1.14 | 0.77 | 1.70 | 0.510 |
| Donor Age | 1.00 | 0.99 | 1.02 | 0.851 |
| Donor Ethnicity/Race (White) | 1.14 | 0.81 | 1.60 | 0.453 |
| Donor Serum Creatinine | 0.94 | 0.73 | 1.22 | 0.657 |
| Center Type (Group B) | 1.36 | 0.96 | 1.92 | 0.084 |

\* Donor Type (DCD vs. DBD), LVAD=Left Ventricular Assist Device, IABP= Intra-Aortic Balloon Pump, Center Type (Group A; centers performed both DCD and DBD SHKT vs. Group B; centers performed only DBD SHKT)

### Sub-group Analysis of Organ Recovery Methods in DCD-SHKT: DPP vs NRP

In the DCD group (N=91), 51 patients had DPP, and 32 patients received NRP. Eight patients were not classified to any group due to missing the time of brain death. Donors in the NRP group were older (DPP, 28 [21-34] vs. NRP, 33 [28.5-39], p=0.002), more likely male (76% vs. 97%, p=0.013), and had higher serum creatinine levels (0.7 [0.6-0.9] vs 0.8 [0.7-1] mg/dl, p=0.007) compared to the DPP group **(Supplementary Table S1)**. The distance of procurement was shorter in the NRP group (400 [188-619] vs. 222.5 [59.3-392.3] miles, p=0.021). Recipients’ EPTS and donor KDPI was higher in the NRP group. Although serum creatinine level at the time of discharge was higher in the DPP group, there was no difference in the rate of KDGF, requirement of dialysis, and kidney graft survival at 6 months (DPP, 90% vs. NRP, 92.6%, p=0.99) during a median follow-up of 256 [151-385.5] days **(Table 4)**. One-year overall recipient (p=0.44) and graft survival (heart p=0.44, kidney p=0.94) were comparable between groups **(Figure 2)**.

**Figure 2.**
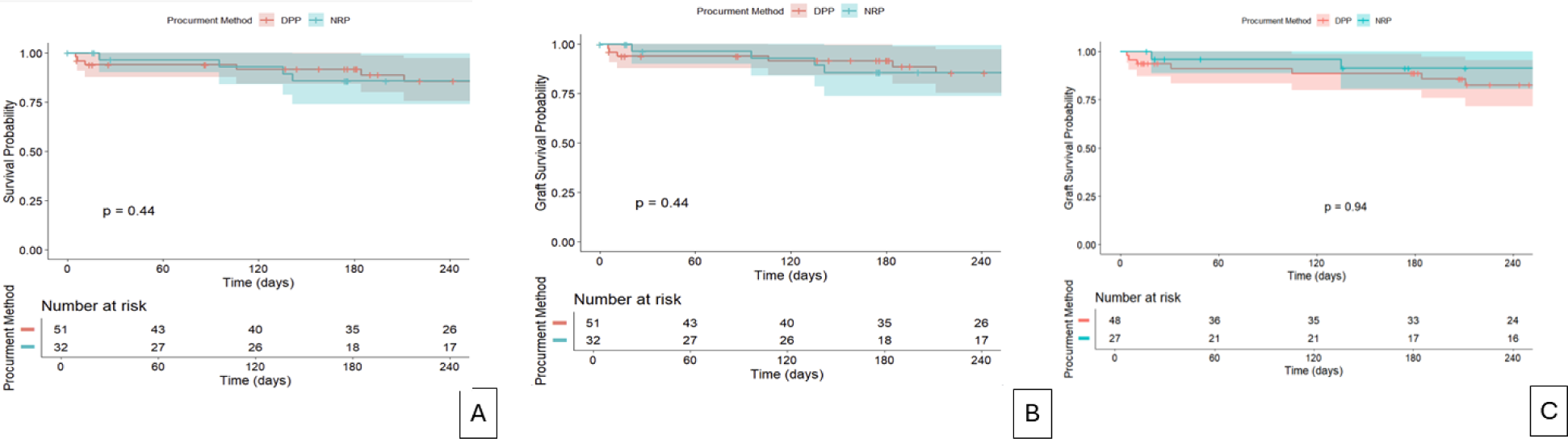
Recipient and Graft survivals outcome in Sub-analysis of DCD cohort by Procurement Method. Direct procurement and perfusion (DPP), Normothermic regional perfusion (NRP), A; Recipient survival, B; Heart graft survival, and C; Kidney graft survival

**Table 4.**
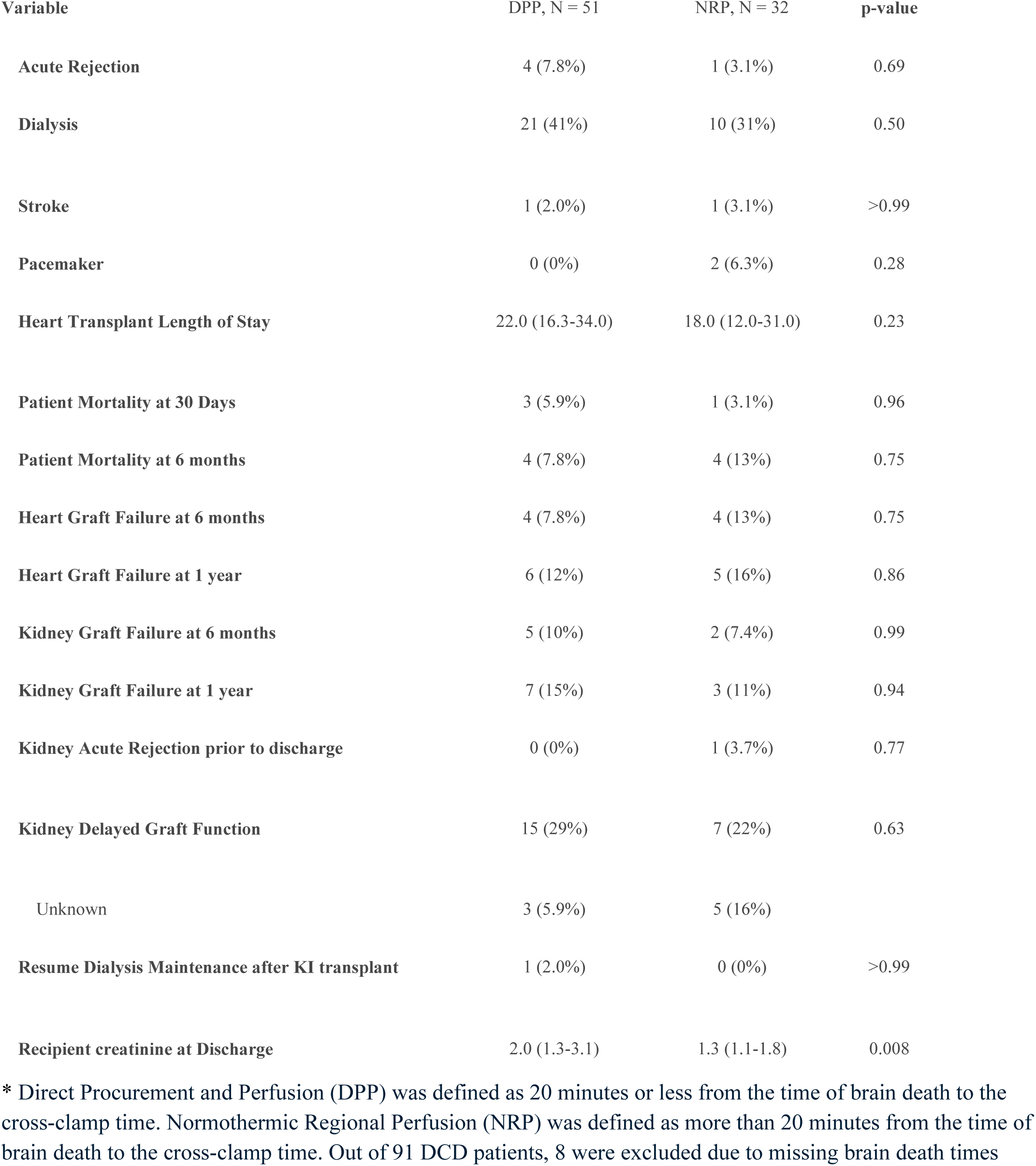
Transplant Outcomes of DCD-SHKT by Recovery Method in non-matched Patients: DPP vs. NRP.

### Sub-group Analysis of Transplant Centers Performing Both DCD- and DBD-SKHT

During the study period, 51.6% of all SHKTs (N=583/1,129) were performed at 30 centers performing both DCD- and DBD-SKHT. Of note, DCD-SHKT accounted for 15.6% (N=91/583) of their SHKT cases (Figure 3). Most centers only performed 1 case of DCD-SHKT while some centers which performed 4 cases or greater showed a high proportion DCD-SHKT per all SHKTs performed at the center **(Figure 3B, Supplementary Table S2)**. Each UNOS region has at least one center which had performed both DCD- and DBD-SHKT **(Figure 3A)**. Region 5 and region 11 performed 24% and 16% of total national DBD-SHKTs and 20% and 27% of national DCD-SHKTs, respectively.

**Figure 3A.**
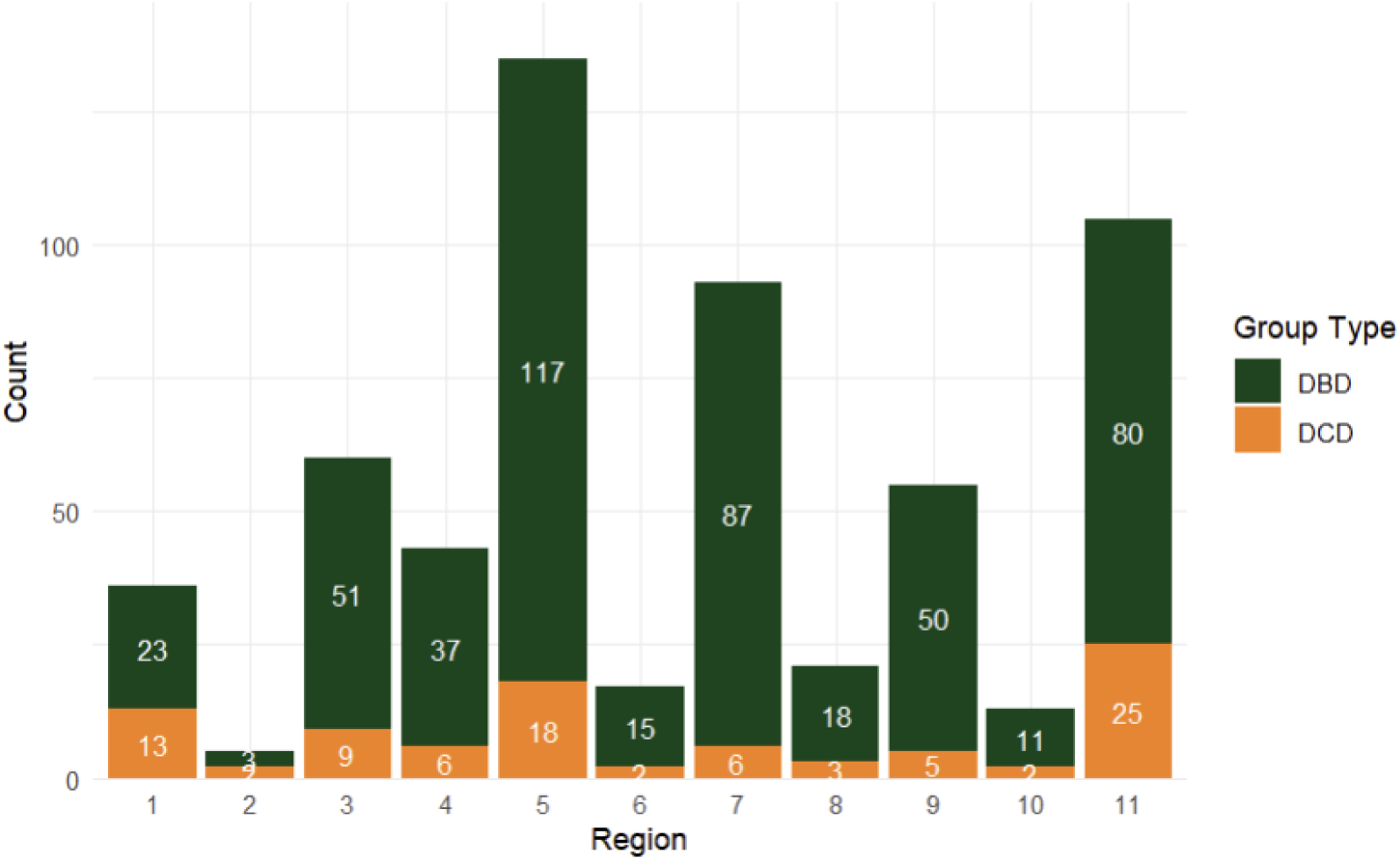
SHKT Cases at Transplant Centers Performing Both DCD and DBD SKHT. **A. Regional DBD and DCD cases at Centers Performing Both DCD and DBD SKHT** UNOS Regions; Connecticut, Eastern Vermont, Maine, Massachusetts, New Hampshire, and Rhode Island, 2; Delaware, District of Columbia, Maryland, New Jersey, Pennsylvania, and West Virginia, 3; Alabama, Arkansas, Florida, Georgia, Louisiana, Mississippi, and Puerto Rico, 4; Oklahoma and Texas, 5; Arizona, California, Nevada, New Mexico, and Utah, 6; Alaska, Hawaii, Idaho, Montana, Oregon, and Washington, 7; Illinois, Minnesota, North Dakota, South Dakota, and Wisconsin, 8; Colorado, Iowa, Kansas, Missouri, Nebraska, and Wyoming, 9; New York and Western Vermont, 10; Indiana, Michigan, and Ohio, 11; Kentucky, North Carolina, South Carolina, Tennessee, and Virginia (https://unos.org/community/regions/)

**Figure 3B.**
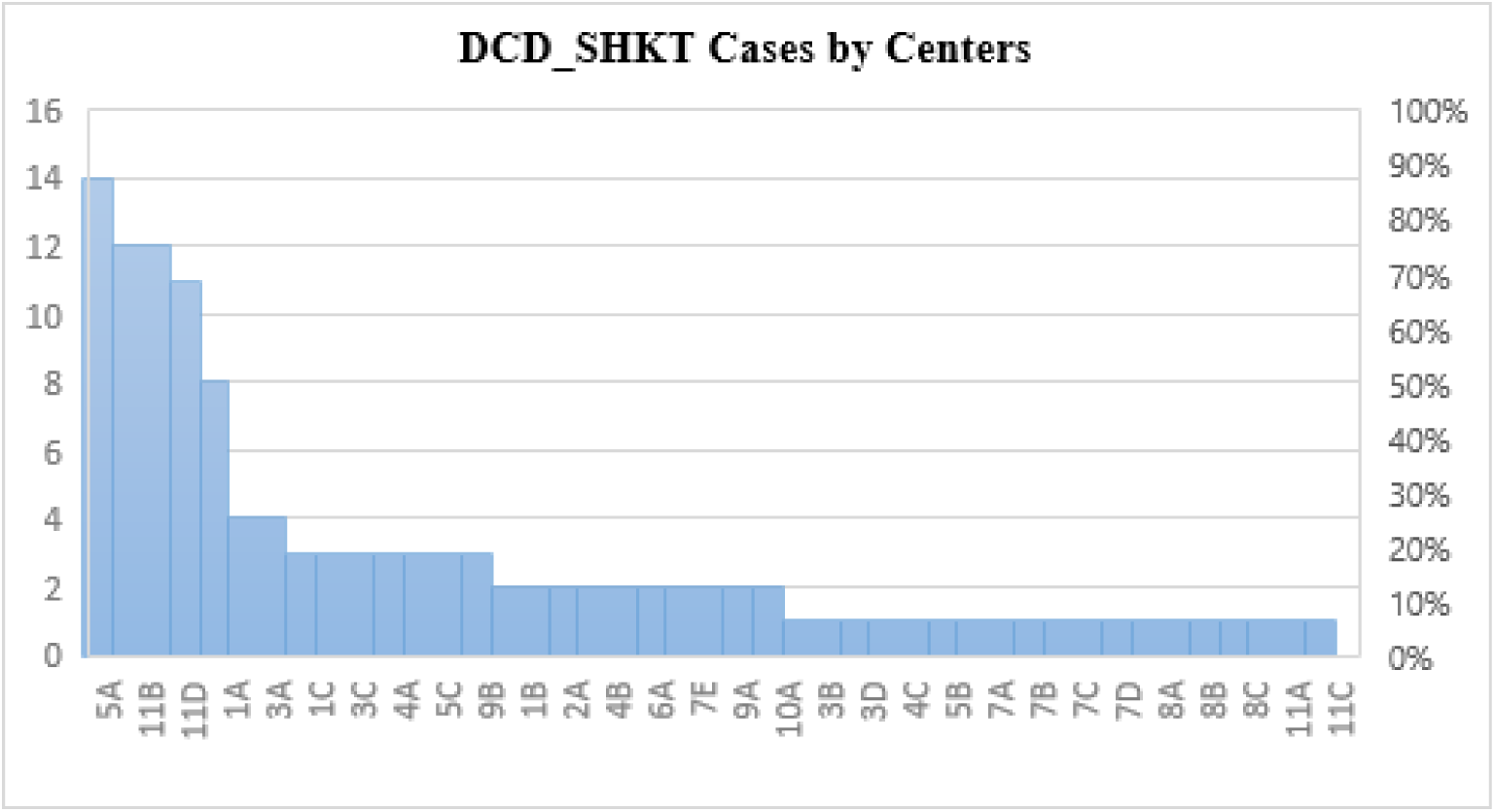
SHKT Cases at Transplant Centers Performing Both DCD and DBD SKHT. **DCD_SHKT Cases by Centers:** Encrypted Center Code was replaced by region and alphabet (e.g., region 1 center A – 1A, region 3 center C – 3C), also shown in Supplementary Table S2.

Among centers performing both DCD- and DBD-SHKT, recipient age was not significantly different but showed a trend towards being older in the DCD group (DCD, 60 [53-65.6] vs. DBD, 58 [50.8-64] years, p=0.054) **(Table 5)**. The usage of durable left ventricular assist device (LVAD) was higher in the DCD group (19% vs. 9.4%, p=0.014). Dialysis at transplant, infection requiring intravenous antibiotics therapy, UNOS status 1 or 2 at the time of OHT, and the usage of intra-aortic balloon pump (IABP) were higher in DBD. DCD donors were younger (30 [24-35] vs. 33 [25-41] years, p=0.002) with lower serum creatinine (0.7 [0.6-1.0] vs. 0.9 [0.7-1.2] mg/dl, p<0.001). The DCD group had shorter time on the kidney waiting list (37 [10-164] vs. 22 [8-77], p=0.011) and a trend towards lower heart waiting list time (27 [9.0-85.3] vs 42 [9.5-160], p=0.07). The incidence of KDGF was similar (27% vs. 27%, p=0.81), however, recipient serum creatinine at discharge was higher in the DCD group (1.6 [1.2-2.4] vs. 1.3 [0.9-1.9] mg/dL, p<0.001). Recipients’ survival and freedom from graft failure were not different between DCD- and DBD-SHKT although numerically higher one-year mortality in the DCD group **(Supplemental Figure S4)**.

**Table 5.** Recipient, Donor, and Organ Procurement Characteristics and Outcomes of Sub-group Cases at Centers Performed Both DCD and DBD SHKT.

| Variable | DBD-SHKT,<br>N = 492<sup>†</sup> | DCD-SHKT,<br>N = 91<sup>†</sup> | p-value<sup>‡</sup> |
| --- | --- | --- | --- |
| **Baseline characteristics** |  |  |  |
| Age | 58.0 (50.8-64.0) | 60.0 (53.0-65.5) | 0.054 |
| Male | 390 (79%) | 74 (81%) | 0.76 |
| Race |  |  | 0.49 |
| Non-White | 275 (56%) | 55 (60%) |  |
| White | 217 (44%) | 36 (40%) |  |
| Diabetes mellitus | 226 (46%) | 50 (55%) | 0.14 |
| BMI | 27.1 (23.8-30.8) | 27.0 (24.0-31.6) | 0.94 |
| Dialysis at transplant | 196 (40%) | 25 (27%) | 0.034 |
| History of smoking | 209 (42%) | 36 (40%) | 0.69 |
| Symptomatic cerebrovascular at registration | 45 (9.1%) | 8 (8.8%) | >0.99 |
| Infection requiring IV drug therapy | 77 (16%) | 6 (6.6%) | 0.035 |
| Status |  |  | <0.001 |
| Status 1-2 | 333 (68%) | 39 (43%) |  |
| Status 3-6 | 159 (32%) | 52 (57%) |  |
| Primary Diagnosis |  |  | 0.61 |
| Ischemic CM | 168 (34%) | 28 (31%) |  |
| Non-Ischemic CM | 324 (66%) | 63 (69%) |  |
| Durable LVAD | 46 (9.3%) | 17 (19%) | 0.014 |
| IABP | 136 (28%) | 8 (8.8%) | <0.001 |
| Impella | 25 (5.1%) | 1 (1.1%) | 0.16 |
| Ventilator support | 13 (2.6%) | 2 (2.2%) | >0.99 |
| ECMO | 21 (4.3%) | 5 (5.5%) | 0.81 |
| History of cardiac surgery | 165 (34%) | 31 (34%) | >0.99 |
| Initial EPTS score | 35 (22-51) | 41 (28-51) | 0.077 |
| Recipient ABO |  |  | 0.12 |
| A | 201 (41%) | 30 (33%) |  |
| AB | 25 (5.1%) | 4 (4.4%) |  |
| B | 88 (18%) | 12 (13%) |  |
| O | 178 (36%) | 45 (49%) |  |
| Donor age | 33.0 (25.0-41.0) | 30.0 (24.0-35.0) | 0.002 |
| Donor male | 372 (76%) | 76 (84%) | 0.13 |
| Donor race |  |  | 0.025 |
| Non-White | 194 (39%) | 24 (26%) |  |
| White | 298 (61%) | 67 (74%) |  |
| **Donor BMI** | 26.8 (23.6-31.1) | 27.1 (24.3-31.6) | 0.44 |
| **Donor Creatinine** | 0.9 (0.7-1.2) | 0.7 (0.6-1.0) | <0.001 |
| **Donor Cause of Death** |  |  | 0.10 |
| Head Trauma | 200 (41%) | 46 (51%) |  |
| Other | 292 (59%) | 45 (49%) |  |
| **Donor LVEF** | 60.0 (57.0-65.0) | 64.0 (60.0-67.0) | 0.10 |
| **Donor KDPI score** | 20 (9-35) | 22 (13-32) | 0.49 |
| **ABO match** |  |  | 0.50 |
| 1 | 410 (83%) | 79 (87%) |  |
| 2 | 82 (17%) | 12 (13%) |  |
| **Procurement Distance (miles)** | 231.5 (76.8-407.0) | 344.0 (153.5-581.5) | <0.001 |
| **Heart Waiting list Time (days)** | 27.5 (9.0-85.3) | 42.0 (9.5-160.5) | 0.071 |
| **Kidney Waiting list Time (days)** | 22.0 (8.0-77.0) | 37.0 (10.0-164.0) | 0.011 |
| **Heart Ischemic Time (hours)** | 3.5 (2.9-4.0) | 4.9 (3.4-6.4) | <0.001 |
| **Heart Machine Perfusion** | 30 / 492 (6.1%) | 53 / 91 (58%) | <0.001 |
| **Kidney Cold Ischemic Time (hours)** | 14.4 (8.5-20.5) | 19.2 (13.7-24.8) | <0.001 |
| **Transplant Outcomes** |  |  |  |
| **Acute Rejection** | 32 / 492 (6.5%) | 6 / 91 (6.6%) | >0.99 |
| **Dialysis** | 172 / 490 (35%) | 34 / 91 (37%) | 0.77 |
| **Stroke** | 33 / 488 (6.8%) | 2 / 91 (2.2%) | 0.15 |
| **Pacemaker** | 5 / 490 (1.0%) | 2 / 91 (2.2%) | 0.67 |
| **Heart Transplant Length of Stay** | 19.0 (14.0-30.0) | 20.0 (15.0-34.0) | 0.29 |
| **Patient Mortality at 30 Days** | 14 / 492 (2.8%) | 5 / 91 (5.5%) | 0.32 |
| **Patient Mortality at 6 months** | 32 / 492 (6.5%) | 9 / 91 (9.9%) | 0.35 |
| **Heart Graft Failure at 6 months** | 33 / 492 (6.7%) | 9 / 91 (9.9%) | 0.39 |
| **Heart Failure at 1 year** | 40 / 492 (8.1%) | 12 / 91 (13%) | 0.18 |
| **Heart Graft Failure at 6 months** | 38 / 447 (8.5%) | 8 / 81 (9.9%) | 0.85 |
| **Kidney Graft Failure at 1 year** | 48 / 447 (11%) | 11 / 81 (14%) | 0.58 |
| **Kidney Acute Rejection prior to discharge** | 7 / 447 (1.6%) | 1 / 81 (1.2%) | >0.99 |
| **Kidney Status at Transplant** |  |  | >0.99 |
| Active | 439 / 447 (98%) | 79 / 81 (98%) |  |
| Temporarily Inactive | 8 / 447 (1.8%) | 2 / 81 (2.5%) |  |
| **Kidney Side, Lt** | 316 / 447 (71%) | 62 / 81 (77%) | 0.35 |
| **Kidney Delayed Graft Function** | 132 / 492 (27%) | 25 / 91 (27%) | 0.81 |
| Unknown | 45 / 492 (9.1%) | 10 / 91 (11%) |  |
| **Resume Dialysis Maintenance after KI transplant** | 11 / 492 (2.2%) | 1 / 91 (1.1%) | 0.76 |
| **Recipient creatinine at Discharge** | 1.3 (0.9-1.9) | 1.6 (1.2-2.4) | <0.001 |

### Sub-group analysis of Transplant Centers Performing Both DCD- and DBD-SKHT vs. Centers Performing only DBD-SKHT

Transplant outcomes of centers that performed both DCD- and DBD-SKHT (Group A) and centers that performed only DBD-SHKT cases (Group B) were compared. There were no differences in baseline characteristics except for a higher percentage of status 3-6 (Group A: 36% vs. Group B: 23%, p=<0.001), lower usage of durable LVAD (11% vs. 17%, p=0.004) and Impella (4.5% vs. 8.2%, p=0.013), and a higher KDPI score (21 [9-35] vs. 16 [7-28] %, p<0.001) in Group A **(Supplementary Table S3)**. Days on the heart waiting list (29 [9-90.5] vs. 39 [14-144.3] days, p<0.001) and kidney waiting list (24 [8-87.3] vs. 35 [12-111] days, p<0.001) were significantly shorter in Group A. Ischemic time was similar (3.5 [3-4] vs. 3.5 [2.9-4.1] hours, p=0.09).

Acute rejection of the heart (Group A: 6.5% vs. Group B: 10%, p=0.024) and length of stay (19 [14-31] vs. 24 [17-36] days, p<0.001) were significantly lower in Group A **(Supplementary Table S3)**. However, the incidence of stroke was higher in Group A (6% vs. 2%, p<0.001). The incidence of KDGF was not different between groups (27% vs. 26%, p=0.54). One-year recipient survival was significantly higher in Group A (91.3% vs. 87%, p=0.043) however freedom from heart (91.1% vs. 87%, p=0.055) and kidney (89% vs. 85%, p=0.10) graft failure were not different **(Supplementary Figure S5)**. In multivariate Cox proportional-hazards analysis, the center Group B was not significantly associated with the higher risk of recipient mortality (HR= 1.35, 95% Cl 0.96-1.91, p=0.088) (**Supplementary Table S4**).

## Discussion

Our comprehensive study found three major findings: Firstly, DCD-SHKT demonstrated comparable survival and graft outcomes compared to DBD-SHKT. Secondly, transplant outcomes in DCD-SHKT were comparable between different modes of organ recovery (DPP vs. NRP). Finally, there was regional and institutional variation in DCD-SHKT volume, and waitlist times were significantly shorter at centers performing both DCD- and DBD-SHKT compared to centers that performed only DBD-SHKT during the study period, without affecting transplant outcomes.

### Overall outcomes of DCD-SHKT

The improved outcomes after isolated DCD transplants have led to utilization of DCD donors for multi-organ transplants. Further, given the number of DBD transplants has plateaued recently^14^, DCD donors could play a crucial role in not only improving waitlist outcomes but also increasing opportunities for candidates with expected prolonged waiting time, such as those with durable LVAD, blood type O, or large BMI patients. We observed some differences in baseline characteristics between DCD-SHKT and DBD-SHKT in the non-matched cohort. This is consistent to the prior reports of isolated DCD heart transplants, which showed that the DCD groups had more patients with status 3-6, a numerically higher rate of durable LVAD, and more blood type O recipients^15,16^. The comparable transplant outcomes between DCD-SHKT and DBD-SHKT shown in this study are notably important given the increasing waitlist times, even for high priority (status 2) patients who constitute the majority under the current allocation policy^17^. Overall, the primary analysis supports the use of DCD donors to enhance waitlist outcomes without negatively impacting overall transplant outcomes.

In general, for SHKT, most centers prefer younger donors, resulting in a lower KDPI (around 20%), which is significantly higher than that of isolated kidney transplant^2,18^. In our study, the KDPI was higher in the DCD group despite the younger donor age. This could be due to the fact that “DCD donor” is one of the variables used to calculate the KDPI, thereby increasing KDPI itself. While the creatinine level at discharge was higher in DCD recipients, possible reflective of impact of warm ischemia on visceral organ functions, the rate of KDGF, dialysis and long-term outcomes were comparable.

### Different types of DCD Recovery Methods in SHKT

In the US, both DPP and NRP procurement methods are utilized for DCD recovery. A major concern with NRP is its ethical implications, and hence is not universally accepted. DPP, on the other hand raises concerns regarding the additional warm ischemic time prior to aortic cross-clamping, due to the need for donor blood collection for ex vivo normothermic perfusion. This process can lead to prolonged warm ischemia of the abdominal organs and lungs, potentially leading to worse transplant outcomes or a higher rate of organ discard^19^. Recent studies focusing on DCD heart transplants have indicated that NRP and DPP procurements are associated with similar one-year post-OHT survival rates^20^.

The impact of prolonged warm ischemia is more pronounced in liver grafts, which are sensitive to warm ischemia and may suffer from biliary necrosis^11,21^. While kidneys are more tolerant of ischemia compared to liver grafts^21–23^, additional nephrectomy time can increase the risk of graft loss in kidneys from DCD donors. In our sub-group analysis, serum creatinine levels at discharge were higher in the DPP group than the NRP group, which might be explained by the difference of abdominal organ recovery between DPP and NRP^11,21^. However, the rate of KDGF, long term need for dialysis and graft survival were not impacted by mode of recovery.

### Transplant Centers performing DCD-SHKT

We found that the centers performing DCD-SHKT accounted for more than half of national SHKT volumes. Although there was regional variation, every state in the US had performed DCD-SHKT, indicating that this emerging recovery method is available to transplant candidates across regions. Among these centers performing both DCD- and DBD-SHKT, a higher number of status 3-6 patients received DCD-SHKT, which correlated with a higher rate of durable LVAD use and blood type O recipients **(Supplemental Table 3)**. This suggests that DCD donors are being effectively utilized to improve the gap between organ demand and donor supply in those centers without negatively impacting outcomes.

Although statistically not significant, survival in patients who received the DCD-SHKT in those DCD centers was lower than DBD-SHKT performed at their centers **(Figure S4)**. This could be the difference of recipients’ characteristics such as recipient age, type of pre-transplant mechanical circulatory support, UNOS status. Finally, centers performing both DCD- and DBD-SHKT demonstrated significantly shorter waitlist times with similar outcomes compared to centers that only performed DBD-SHKT. Previous studies reported that patients who spend double the time on the waitlist have a 10% higher chance of transplant failure^24^ and a higher risk of 90-day mortality if they wait more than eight days on the waitlist, among patients supported with pre-transplant veno-arterial extracorporeal membrane oxygenation (VA-ECMO)^25^. In this regard, SHKT utilizing organs from DCD donors can potentially reduce the days of waitlist and improve waiting list mortality without affecting transplant outcome in patients with end-stage heart failure with kidney dysfunction who otherwise may have to wait longer until DBD donors become available^26^.

### Limitations

Several limitations should be noted. First, this study is retrospective and relies on national registry data, which means that unmeasured variables in the UNOS database could influence the outcomes. Additionally, the follow-up period for the DCD group is limited due to the relatively short history of DCD heart transplants in the US. Outcomes may differ with a larger sample size and a longer follow-up period. It is also possible that more transplant centers are currently performing both DCD- and DBD-SHKT. Also, the database lacks variables to distinguish the duration of pre-transplant dialysis in HKT recipients. Finally, early cardiac graft outcomes (e.g., requirement of VA-ECMO, rate of severe primary graft dysfunction [PGD], etc.) were not available during the study period. While several studies have shown that a higher incidence of severe PGD in DCD heart transplant does not affect one-year survival compared to DBD, little is known about how severe PGD might impact subsequent kidney transplants in multi-organ DCD transplants. There is also a possibility that a subsequent kidney transplant might be aborted if severe PGD occurs in the cardiac graft. Granular analysis using the PGD data in UNOS database will hopefully provide more insight on early outcome of SHKT or isolated OHT in patients with kidney dysfunction.

## Conclusion

Early to mid-term outcomes of SHKT utilizing DCD donors yield comparable survival and graft outcomes to those from DBD donors. In DCD-SHKT, there is no significant difference in survival or transplant outcomes between donors recovered using DPP or NRP. Moreover, centers performing both DCD- and DBD-SHKT accounted for over 50% of the national SHKT volume. Our study suggests that utilizing DCD donors for SHKT can safely expand the donor pool without compromising transplant outcomes. These findings will serve as a benchmark for donor/recipient selection and transplant strategies, including the indication of multi-organ transplants using DCD donors as well as the implementation of a kidney safety net policy in candidates with kidney dysfunction.

## Data Availability

All data related to in the manuscript is availble at United Network for Organ Sharing.

## Acknowledgments

None

